# Pathways linking physical and mental health: The role of brain structure and environment

**DOI:** 10.1101/2024.01.15.24301190

**Authors:** Ye Ella Tian, James H Cole, Edward T Bullmore, Andrew Zalesky

**Affiliations:** Department of Psychiatry, Melbourne Medical School, The University of Melbourne; Melbourne, Australia; Centre for Medical Image Computing, Department of Computer Science, University College London, UK; Dementia Research Centre, Queen Square Institute of Neurology, University College London, UK; Department of Psychiatry, University of Cambridge, Cambridge, UK; Cambridgeshire & Peterborough NHS Foundation Trust, Cambridge, UK; Department of Biomedical Engineering, Faculty of Engineering and Information Technology, The University of Melbourne; Melbourne, Australia

## Abstract

Depression and anxiety are prevalent in people with a chronic physical illness. Increasing evidence suggests that co-occurring physical and mental illness is associated with shared biological pathways. However, little is known about the brain’s role in mediating links between physical and mental health. Using multimodal brain imaging and organ-specific physiological markers from the UK Biobank, we establish prospective associations between the baseline health of seven body organs and mental health outcomes at 4-14 years follow-up, focussing on depression and anxiety. We reveal multiple pathways, mediated by the brain, through which poor organ health may lead to poor mental health. We identify several lifestyle factors that influence mental health through their selective impact on the physiology of specific organ systems and brain structure. Our work reveals the interplay between brain, body and lifestyle and their collective influence on mental health. Pathways elucidated here may inform behavioral interventions to mitigate or prevent the synergistic co-occurrence of physical and mental disorders.

## Introduction

Depression and anxiety are common mental health problems with shared neurobiology ^1,2^ and genetic risk ^3^. They are not only highly co-occurrent ^4^, the symptoms of depression and/or anxiety also frequently manifest in individuals primarily diagnosed with other mental disorders such as schizophrenia ^5^ and bipolar disorder ^6^. The presence of co-occurring depression and anxiety often complicates the course of mental illness and leads to greater functional impairment and poorer clinical outcomes ^6,7^.

The prevalence of depression and anxiety is also high in individuals with a chronic physical health conditions ^8,9^, and the risk of developing depression and anxiety is several-fold higher in these patients compared to people without medical disorders ^8,10,11^. Indeed, an emerging body of evidence suggests that chronic physical illness, including coronary heart disease, respiratory disease, diabetes, musculoskeletal disorders and cancer may lead to the development of, or prolonged mental health problems, particularly depression and anxiety in middle-aged and older adults ^12–15^. Comorbid depression and anxiety impact an individual’s overall health and associate with worse prognosis, poorer quality of life and increased mortality ^8^.

Multiple putative pathways through which physical illness leads to, or exacerbates, depressive and anxiety symptoms have been proposed in the literature, including but not limited to i) converging biological pathways, such as shared genetic disposition ^16,17^, immunometabolic dysregulation ^18^ and pain-related alterations in monoamine levels ^19^; ii) behavioral and lifestyle factors such as smoking, poor nutrition, lack of physical activity and sleep disturbances ^20,21^; and, iii) psychological pathways such as emotional distress and the fear of impending death due to chronic illness ^22^.

These proposed pathways linking physical and mental health are supported by epidemiological studies; however, they remain poorly understood because the complex interplay between these mechanisms could co-occur and underlie the development of physical-mental comorbidity. Importantly, the brain is rarely considered in these models, despite extensive evidence of changes in brain structure and function associated with depression and anxiety ^23–26^. Further work is needed to understand the potentially causal or mediating role of the brain on the concurrent manifestation of physical and mental illness. This knowledge may provide impetus to alleviate the mental-physical dichotomy in medicine and facilitate integrated and holistic health care across psychiatry and other medical disciplines.

The brain is intricately interconnected with other organ systems via multiple biophysiological axes such as brain-heart, brain-lung, brain-liver and brain-endocrine systems ^27–30^. The health and function of the brain can hence be influenced by other organ systems. For example, a recent study using longitudinal data showed that premature aging of some organ systems (e.g., cardiovascular) at middle age is associated with an older-appearing brain later in life ^31^. This suggests that poor physical health may lead to poor brain health. As such, the health and function of the brain may mediate the relationship between physical and mental health.

Most previous studies investigating the relationship between physical and mental health are epidemiological and focus on comorbidity rates among major medical conditions and clinically diagnosed major depressive disorder and/or anxiety disorders. However, our recent study assesses physiological markers and provides biological evidence suggesting a largely shared imprint of poor physical health across common mental illnesses ^32^. Moreover, subthreshold depressive and anxiety conditions are prevalent in the general population ^33,34^, and the potential consequences on the physical health in such individuals is unknown. Work is needed to understand the dynamic and dose-dependent relationship ^14^ between physical and mental health, and the role of the brain in potentially mediating this relationship.

In this study, we use multimodal brain imaging, physiological, blood- and urine-derived markers from the UK Biobank ^35,36^, the world-largest prospective population-based cohort, to systematically investigate the extent to which the health and function of multiple organ systems associates with depression and anxiety symptoms. Although our recent study ^32^ suggests markedly poorer physical health in people with common mental disorders compared to their age- and sex-matched healthy peers, the relationship between physical health and mental health symptom severity was not investigated. We sought to assess whether this relationship is mediated by individual differences in brain structure and identify modifiable lifestyle factors that can potentially improve physical and brain health and consequently alleviate depression and anxiety symptoms. Our work provides a consolidated view of pathways explaining the interplay between brain, body and environment and how they collectively influence mental health outcomes.

## Results

A subset of UK Biobank participants (n=18,083, 7,433 males) was studied. This subset comprises 7,749 individuals (3,774 males) without any clinically diagnosed major medical and mental conditions, and 10,334 individuals (3,659 males) who reported a lifetime diagnosis of one or more of the following four mental disorders: schizophrenia (n=67), bipolar disorder (n=592), depression (n=9,817) and generalized anxiety disorder (n=2,041). Baseline demographic characteristics is provided in Supplementary Table 1.

Physical health was separately assessed for 7 organ systems (cardiovascular, pulmonary, musculoskeletal, immune, renal, hepatic and metabolic) using organ-specific phenotypes for each individual (age range: 40-70 years, mean 53.7 ± 7.3), as described previously ^32^. Specifically, the health status of each organ was indexed using a composite organ health score, which quantifies the extent to which an individual’s organ health and function deviates from an age- and sex-specific normative reference ranges (Figure 1a). Organ health scores were calibrated such that a score of zero indicated normal organ function (population median), whereas a negative value suggested compromised organ health, controlling for age and sex (Methods).

**Figure 1.**
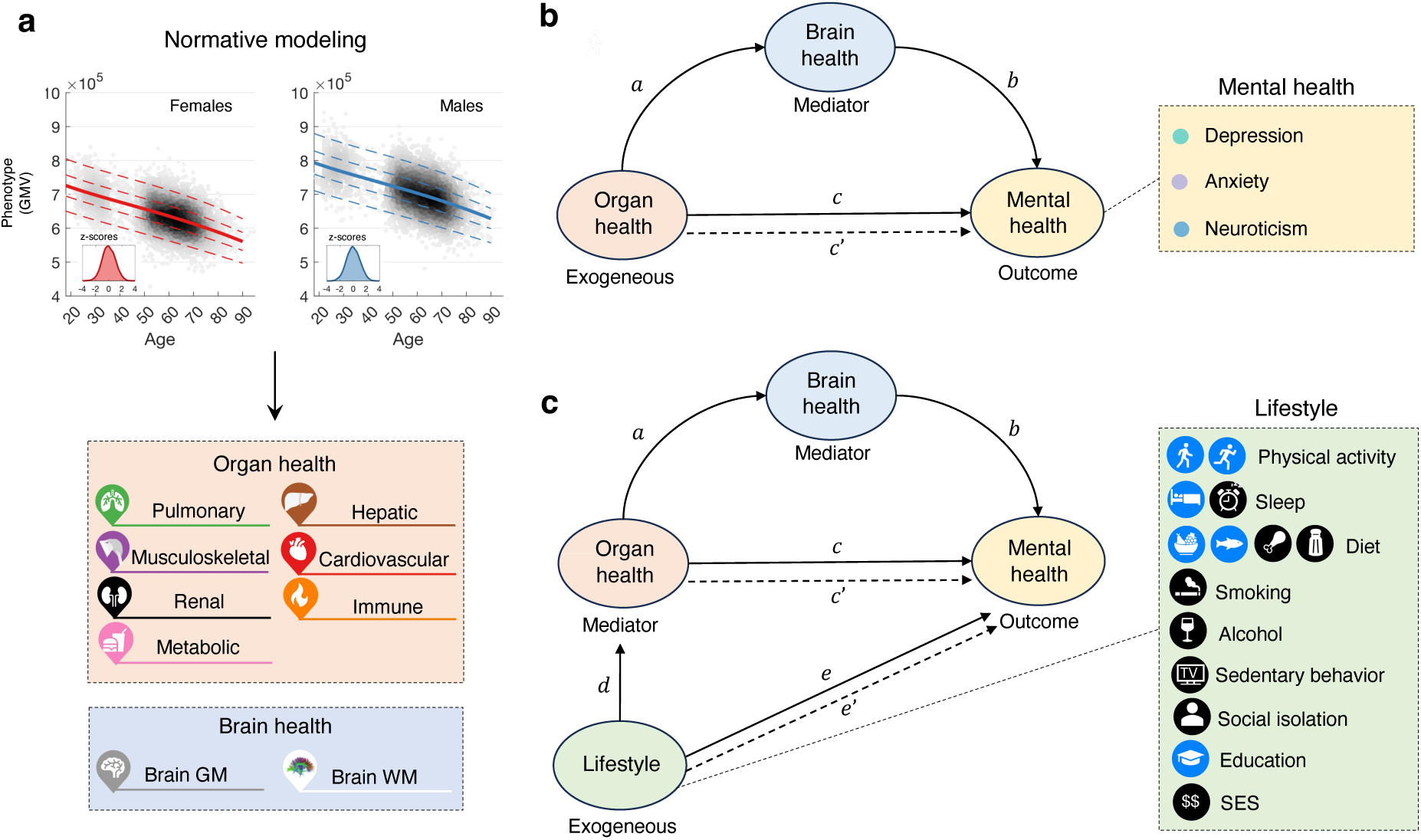
Overview of study design. **a**), Schematic of organ health score estimation. Normative models of age-related changes in each physiological and brain imaging-derived phenotype were established using generalized additive models for location, scale and shape in a group of healthy individuals, stratified by sex. The median is denoted by a solid line and the dotted lines indicate 5%, 25%, 75% and 95% centiles (from bottom to top). The inset shows the distribution of the standardized phenotypic deviation scores (z-scores) across individuals. An organ health score was computed for each organ system and for each individual using organ-specific phenotypic deviation scores. **b**), Schematic of structural equation model (SEM, path analysis) formulated to test whether brain health mediates the association between organ and mental health (depression, anxiety and neuroticism). The mediation effect is deemed significant if the product of regression coefficients *a*×*b* is significant under the condition of a significant total effect *c’* and a satisfactory goodness of model fit. **c**), Schematic of SEM formulated to identify lifestyle factors that influence mental health outcomes through influencing organ health and brain health. The mediation effect is deemed significant if the product of regression coefficients *a*×*b*×*d* is significant under the condition of a significant total effect *e*’ and a satisfactory goodness of model fit. GM, gray matter; WM, white matter; SES, socioeconomical status indexed by the Townsend deprivation index.

Brain imaging data, including structural and diffusion-weighted magnetic resonance imaging (MRI), were acquired at 4-14 years following the assessment of organ health as part of follow-up visits for these individuals (age range: 45-83 years, mean 62.7 ± 7.5). For each individual, a standardized phenotypic deviation score was calculated for the volume of each brain gray matter (GM) region, and the mean fractional anisotropy (FA) of each white matter (WM) tract based on established normative models of brain structure described previously (Figure 1a) ^32^. A deviation score quantifies the number of standard deviations an individual deviated from the normative reference median ^37^.

Depressive symptom severity was assessed using the Recent Depressive Symptoms (RDS-4) scale ^38^ on the day of brain scanning. Anxiety symptom severity was assessed using the Generalized Anxiety Disorder (GAD-7) scale at a separate online follow-up ^39^. The time difference between the online questionnaire and brain scan ranges from -1303 to 989 days (median: -513 ± 550, negative values indicate assessment completed prior to scanning). To complement the time discrepancy, the Eysenck Neuroticism (N-12) score assessed on the day of brain scanning was also considered due to its known association with the development of depression and anxiety ^40^. The severity of depressive and anxiety symptoms and neuroticism varies widely across the 5 diagnosis groups. Individuals with generalized anxiety disorder on average manifest the highest score across all three mental health domains and healthy comparison individuals manifest the lowest. The presence of depressive and anxiety symptoms in the healthy comparison group may indicate subthreshold depressive and/or anxiety disorder (Supplementary Figure 1). As such, our analyses focused on the pool of all individuals, irrespective of their lifetime diagnosis.

Given the moderate interindividual associations in symptom measures (Pearson correlation coefficient range: 0.51-0.58), our core analyses were repeated for a general summary measure of mental health status derived using principal component analysis (Methods).

### Physical health of multiple organ systems associates with mental health

A prospective association was evident between baseline physical health and mental health outcomes at follow-up. For each of the 7 organ systems, we found that poorer organ health was significantly associated with higher depressive symptoms, controlling for age at physical health assessment and sex (β range: -0.090 to -0.039, *p*<0.05, two-tailed, false discovery rate (FDR) corrected across 7 organs; Supplementary Figure 2a). Similarly, poor organ health scores, except renal and pulmonary system scores, were significantly associated with higher anxiety symptoms (β range: -0.085 to -0.037, *p*<0.05, two-tailed, FDR corrected across 7 organs, Supplementary Figure 2b). Poorer health scores of all organs, except the renal system, significantly associated with higher neuroticism (β range: -0.104 to -0.041, *p*<0.05, two-tailed, FDR corrected across 7 organs, Supplementary Figure 2c). The musculoskeletal system consistently showed the strongest associations with the three mental health measures, followed by the immune, metabolic and hepatic systems. For each standard deviation reduction in musculoskeletal health, an individual can expect an approximately 0.1 standard deviation worsening of mental health symptoms. Physical-mental health associations were modest for cardiovascular and pulmonary systems and the weakest for the renal system.

### Brain health mediates the link between physical and mental health

We next investigated whether the prospective association between physical (organ) and mental health is mediated by the brain. To this end, a structural equation model (SEM, path analysis), was formulated for each organ-mental health pair showing a significant association (Supplementary Figure 2), with the brain as the mediator, mental health as the outcome variable and organ health as the exogeneous factor. Of note, the relationship between physical and mental health is likely bi-directional and state-dependent ^41^; mental illness could also lead to subsequent development of medical conditions affecting various body systems ^42^. Here, we specifically tested pathways from organ health to mental health outcome due to the chronological nature of participant assessment in the UK Biobank (i.e., physical health predates brain imaging and mental health assessments).

We tested the significance of the mediating effect (*a*×*b*, Figure 1b) of brain GM and WM, respectively. We first analyzed deviation scores of global brain measures, including the total GM volume and the mean FA across all WM tracts. Regional analyses were conducted if global brain measures showed significant mediating effects. Overall, we found multiple significant pathways through which poor organ health may lead to poor brain health, which in turns lead to poor mental health (Figures 2&3). The extent to which brain structure mediates physical-mental health varies across organ systems. In general, the brain showed a strong mediating effect on organs that had strong direct effects on mental health outcomes; namely, the musculoskeletal and immune systems. However, pathways were observed linking specific organ systems to mental health symptoms through the brain. This is exemplified by the pulmonary and cardiovascular systems. Specifically, we found that GM showed a significant mediating effect on pulmonary-depression (Figure 2a) and pulmonary-neuroticism (Figure 2c), but not pulmonary-anxiety (Figure 2b) associations; WM showed a significant mediating effect on associations of cardiovascular-anxiety (Figure 3b) and cardiovascular-neuroticism (Figure 3c), but not cardiovascular-depression (Figure 3a). Supplementary analyses using the summary score of mental health outcomes showed consistent findings, in that poor physical health of all organs except the renal system led to poor mental health via changes in brain structure (Supplementary Figure 3)

**Figure 2.**
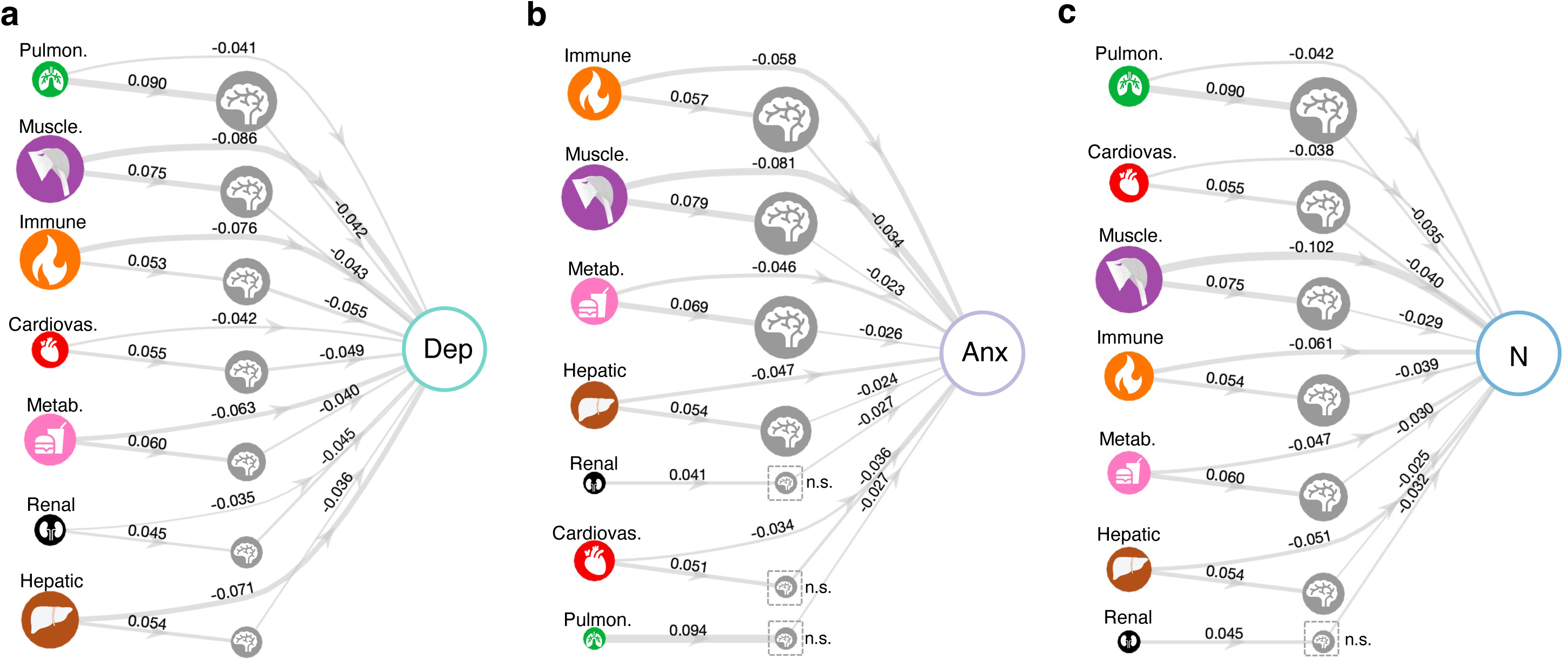
Brain gray matter volume mediates physical-mental health associations. Pathways linking organ health, brain gray matter volume and mental health. A structural equation model (SEM) linking the health of each organ system to the severity of depression (**a**), anxiety (**b**) and neuroticism (**c**) was fitted with brain gray matter volume as the mediating factor. Links are shown for significant paths linking organ health (exogeneous) and mental health (outcome) via brain gray matter volume (mediator) inferred from SEM (false discovery rate corrected for 7 organs). Node size of organs is modulated by the direct effect from organs to mental health outcome. Node size of the brain is modulated by its mediating effect and ranked in decreasing order. Edge thickness reflects regression coefficients estimated for edges comprising the SEM. Non-significant mediating effect of the brain is indicated by a dashed square. Pulmon., pulmonary; Muscle., musculoskeletal; Cardiovas., cardiovascular; Dep, depression; Anx, anxiety; N, neuroticism; n.s., non-significant.

**Figure 3.**
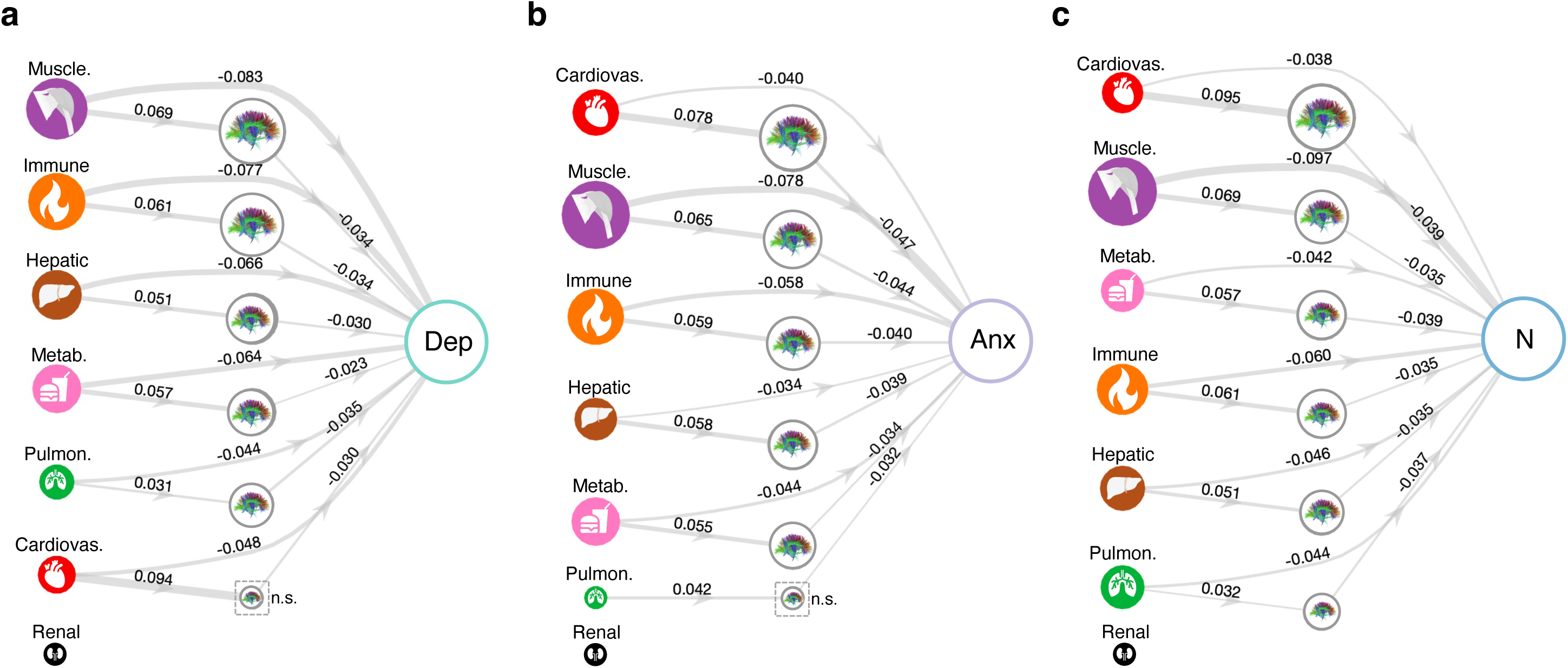
Brain white matter microstructure mediates physical-mental health associations. Pathways linking organ health, fractional anisotropy (FA) of white matter and mental health. A structural equation model (SEM) was fitted for each organ system and for depression (**a**), anxiety (**b**) and neuroticism (**c**), respectively. Links are shown for significant paths linking organ health (exogeneous) and mental health (outcome) via brain white matter (mediator) inferred from SEM (p<0.05, false discovery rate corrected for 7 organs). Node size of organs is modulated by the direct effect from organs to mental health outcome. Node size of the brain is modulated by its mediating effect. Edge thickness reflects regression coefficient estimated for edges comprising the SEM. Non-significant mediating effect of the brain is indicated by a dashed square. Pulmon., pulmonary; Muscle., musculoskeletal; Cardiovas., cardiovascular; Dep, depression; Anx, anxiety; N, neuroticism; n.s., non-significant.

Regional analyses revealed significant involvement of multiple brain GM regions in mediating the relationship between physical health and depression (Figure 4a). Mediation effects were most prominent within temporal, medial frontal, insula and somatomotor areas. Some of these regions were also involved in mediating the impact of organ health on anxiety and neuroticism. For anxiety, the middle temporal, banks of superior temporal sulcus, inferior parietal areas showed the strongest mediating effects (Figure 4b), and medial orbitofrontal, insula and precentral gyrus for neuroticism (Figure 4c). Across the three mental health measures, GM volume imparted the largest impact on depressive symptom severity, and the influence was the weakest on anxiety. Multiple WM tracts mediated the relationship between physical health and depressive symptoms. Fewer WM tracts mediated effects for anxiety and neuroticism. WM tracts with the strongest mediating effects included superior fronto-occipital fasciculus for depression (Figure 4d), anterior corona radiata for anxiety (Figure 4e) and posterior thalamic radiation for neuroticism (Figure 4f). Brain regions that showed significant medicating effects for individual mental health measure were also evident for the summary score of mental health outcomes (Supplementary Figure 4).

**Figure 4.**
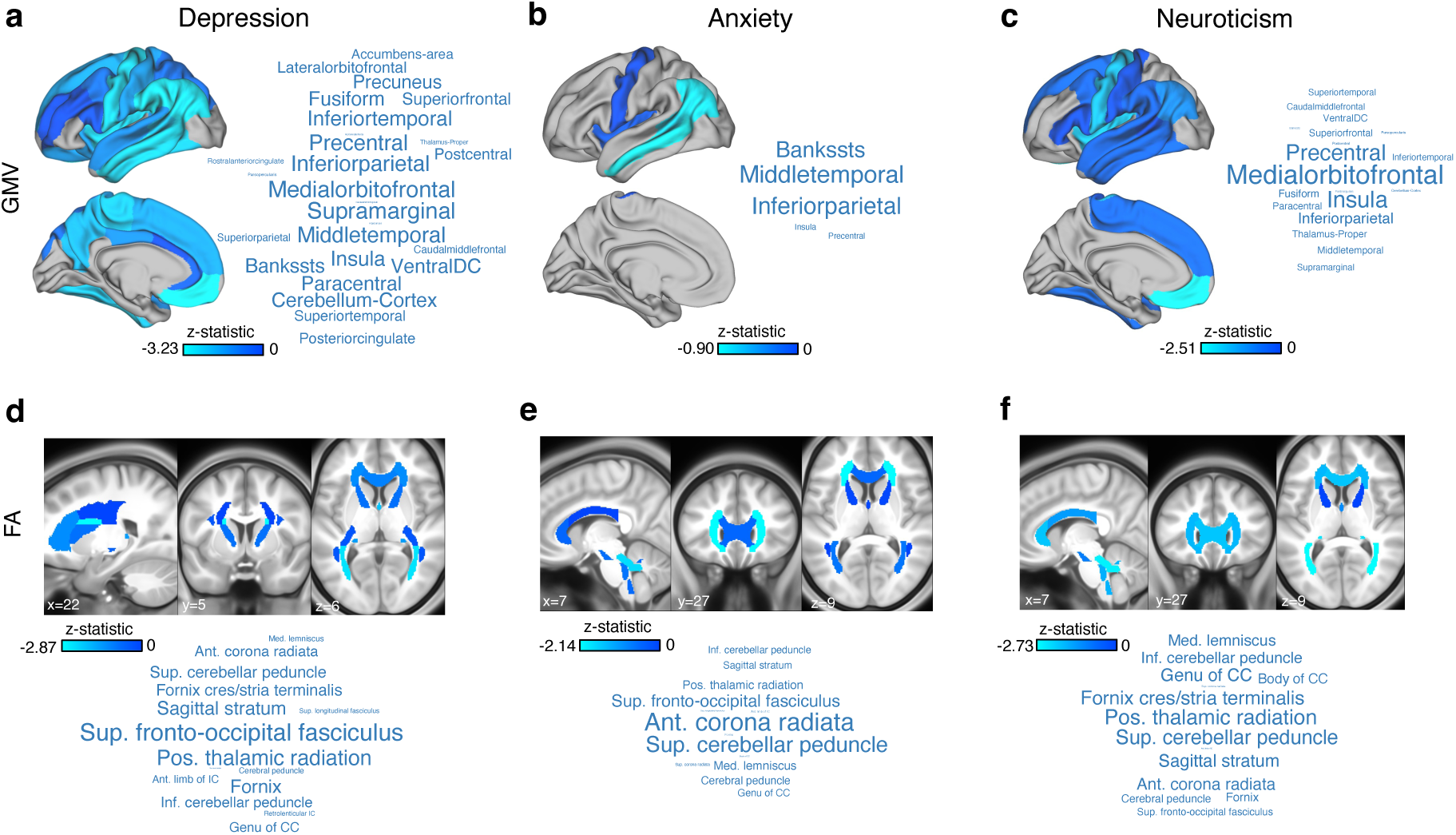
Mediating effects of gray and white matter regions on physical-mental health associations. A structural equation model (SEM) was fitted for each gray matter region, white matter tract and for each organ system. Significant mediating effects (z-statistic) of brain regions (p<0.05, FDR-corrected for 33 gray and 27 white matter regions) were averaged across 7 organ systems to provide a consensus mediating effect map for depression (**a,d**), anxiety (**b,c**) and neuroticism (**c,f**), respectively. The average z-statistic for cortical gray matter volume (Desikan-Killianny atlas) are rendered on cortical surface for visualization. Word clouds show top-ranked cortical and subcortical regions. The font size is scaled according to the absolute value of the average z-statistic. Similarly, the average z-statistic for regional fractional anisotropy (JHU ICBM-DTI-81 atlas) are rendered in standard Montreal Neurological Institute (MNI)-152 anatomical space. Word clouds show top-ranked white matter tracts. The font size is scaled according to the absolute value of the average z-statistic.

### Lifestyle factors influence mental health via physical and brain health mediators

Lifestyle choices including smoking, alcohol use, physical inactivity, poor nutrition and sleep are known risk factors for poor physical health and chronic diseases ^43^. Emerging evidence suggests an impact of lifestyle on mental health and psychological well-being via potentially shared neurobiological pathways ^21,44^. Yet, little is known about the pathways through which lifestyle influences mental health through changes in physical health and neurobiology. We sought to identify lifestyle factors that lead to poor mental health via their impact on organ health and the brain (Figure 1c). We selected 14 lifestyle factors commonly associated with physical and/or mental health. Interestingly, we found that while most lifestyle factors assessed at baseline were significantly associated with all three mental health outcome measures (p<0.05, FDR corrected across 14 lifestyle factors x 3 mental health measures=42 tests, Supplementary Figure 5), alcohol consumption and diet were exceptions. Specifically, we found that more alcohol consumption was associated with higher anxiety and neuroticism scores but not with depression, and that oily fish, fruit and vegetable intake was associated with lower depressive and neuroticism scores but not with lower anxiety symptoms. Among these lifestyle factors, sleep quality (i.e., insomnia, sleep duration) and walking pace consistently showed the strongest associations with the three mental health measures.

Using SEM (path analysis), we found that poor physical and brain health collectively mediated the associations between baseline lifestyle exposure and mental health outcomes of depression and neuroticism. However, the mediation effects were not significant for anxiety. Significant pathways were identified for all 7 organ systems for depression via brain GM, but not via WM (Figure 5). In contrast, significant pathways leading to neuroticism were via WM and were evident for 5 organ systems including immune, musculoskeletal, metabolic, hepatic and pulmonary system (Figure 6). Notably, we found that some lifestyle factors, including physical activity, sedentary behavior, diet, sleep quality, degree of education and socioeconomic inequality, influenced mental health via influencing multiple body and brain systems. In contrast, other lifestyle factors, particularly smoking and alcohol consumption, were involved in more specific paths affecting mental health through the metabolic and pulmonary systems (Figure 5c,f, Figure 6e,f), and musculoskeletal and hepatic systems (Figure 6b, d), respectively. For example, smoking could worsen metabolic and pulmonary health, which in turns leads to reductions in brain gray matter volume and subsequent depressive symptoms. In addition, we found that while a lack of social connection was associated with higher scores of depression, anxiety and neuroticism, this association was not significantly mediated by the physical-brain pathway, which may suggest a direct impact of psychosocial factors on mental health.

**Figure 5.**
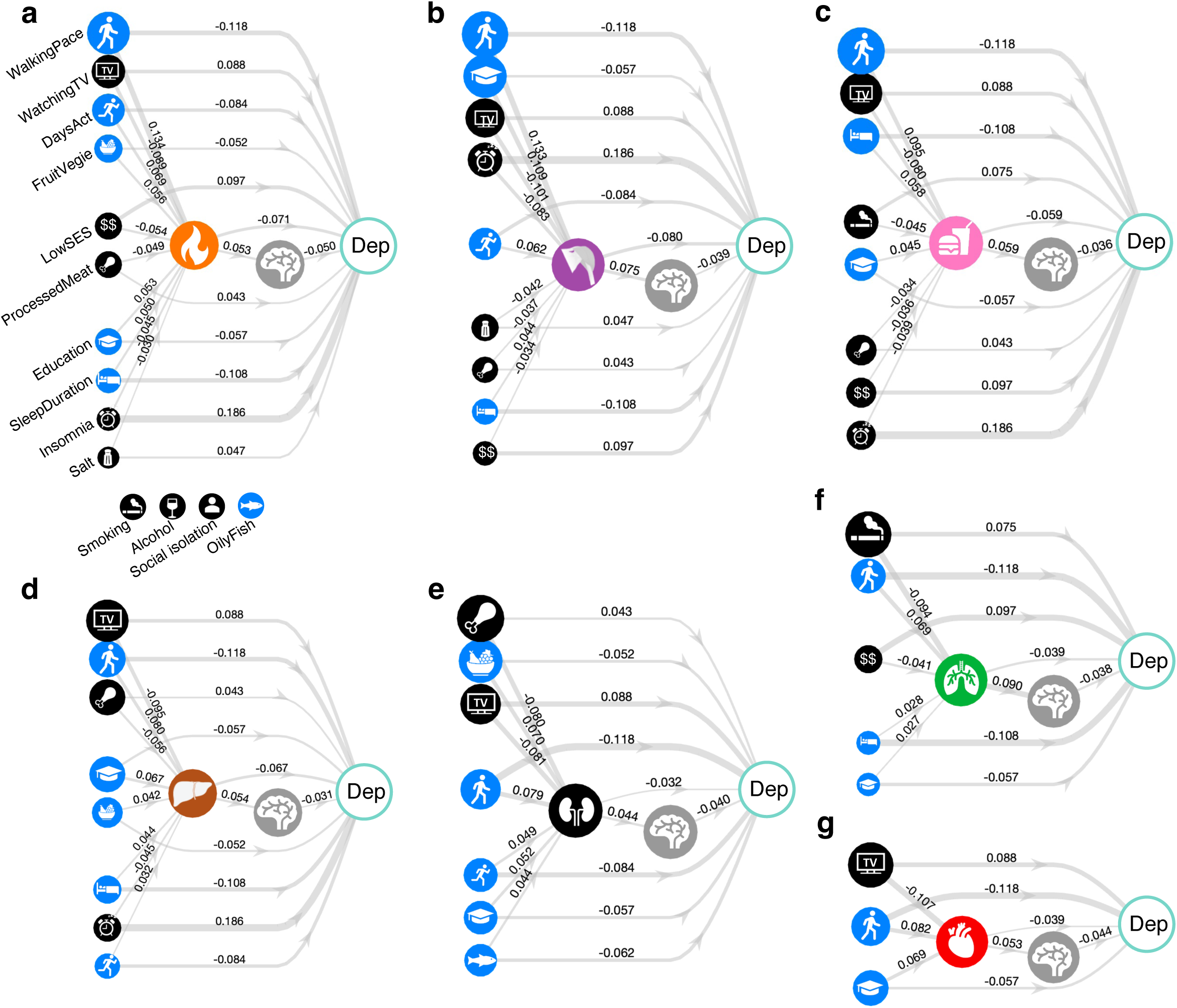
Physical and neurobiological pathways through which lifestyle factors influence depression severity. A structural equation model (SEM) was separately fitted for each lifestyle factor and for each of the 7 body organs, to test whether lifestyle factors influence depression symptom via influencing organ health and brain structure. Links are shown for significant paths where organ health and brain gray matter volume had significant mediating effects (p<0.05, FDR corrected across 14 lifestyle factors). The mean FA of white matter tracts did not show significant mediating effect in this analysis and thus not shown. Node size of lifestyle factors is modulated by the combined mediating effect of the organ and the brain (i.e., *a*×*b*×*d* shown in Figure 1c). Lifestyles factors associated with higher/lower depressive symptom severity are colored black/blue. Edge thickness reflects regression coefficient estimated for edges comprising the SEM. Edges comprising non-significant mediating paths are supressed.

**Figure 6.**
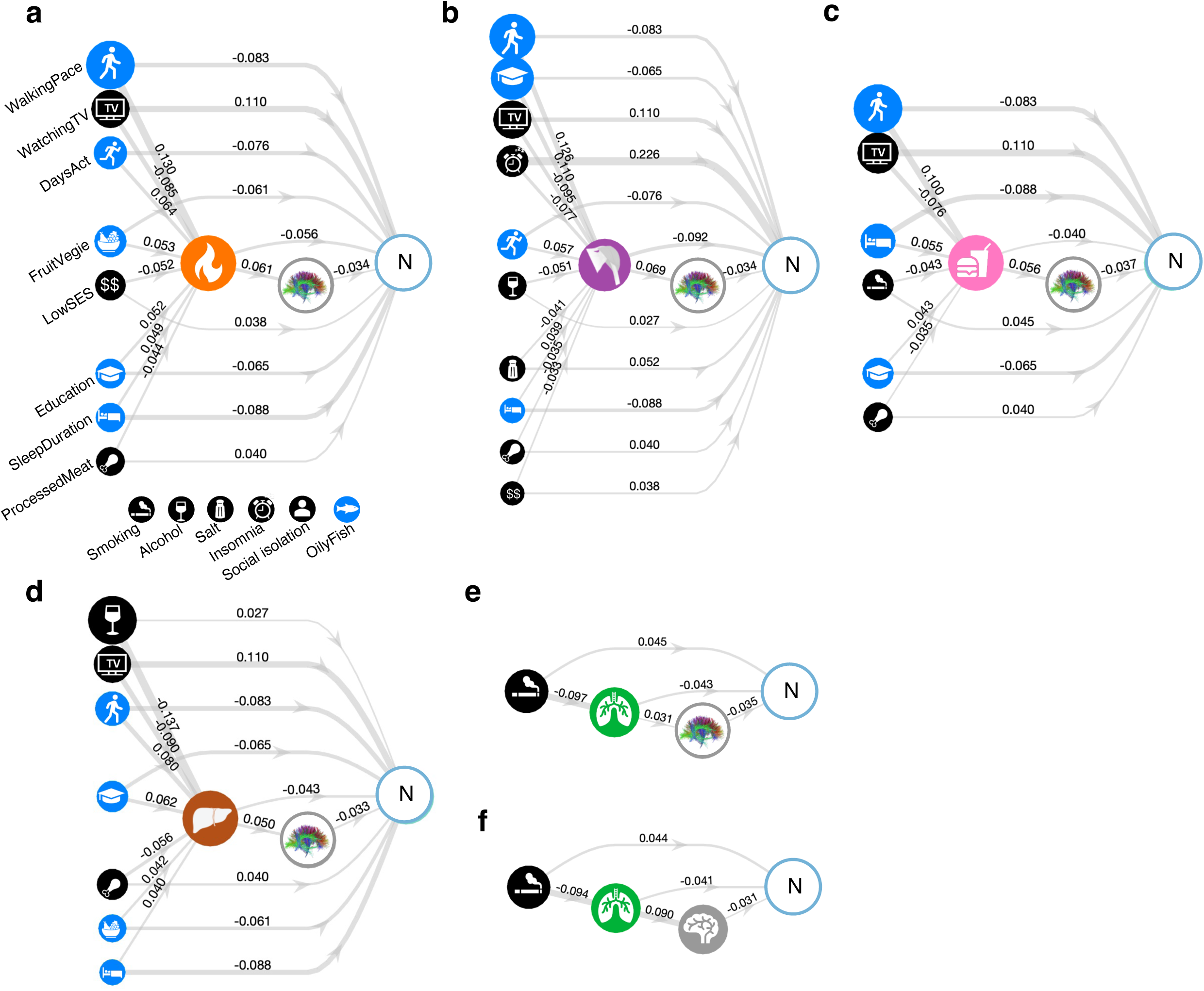
Physical and neurobiological pathways through which lifestyle factors influence neuroticism. A structural equation model (SEM) was separately fitted for each lifestyle factor and for each of the 7 body organs, to test whether lifestyle factor influences neuroticism via influencing organ health and brain structure. Links are shown for significant paths where organ health and brain structure (total gray matter volume or mean FA of white matter tracts) had significant mediating effects (p<0.05, FDR corrected across 14 lifestyle factors). Node size of lifestyle factors is modulated by the combined mediating effect of the organ and the brain (i.e., *a*×*b*×*d* shown in Figure 1c). Lifestyles factors associated with higher/lower neuroticism score are colored black/blue. Edge thickness reflects regression coefficient estimated for edges comprising the SEM. Edges comprising non-significant mediating paths are supressed.

## Discussion

By integrating clinical data, brain imaging and a wide array of organ-specific biomarkers in a large population-based cohort, we elucidated for the first time multiple putative pathways involving the brain as a mediating factor and through which poor physical health of body organ systems leads to poor mental health. We identified modifiable lifestyle factors that can potentially lead to improved mental health through their impact on these specific organ systems and neurobiology. Our work provides a holistic characterization of brain, body, lifestyle and mental health.

Somatic symptoms and physical comorbidity in individuals with depression and/or anxiety disorder have been increasingly linked to immunometabolic dysregulation, e.g., chronic low-grade inflammation and imbalanced energy homeostasis ^18,45^. Our work extends this notion by showing that depression and anxiety are not only associated with dysfunction of immune and metabolic systems, but additionally with poor physical health of multiple other organ systems, particularly of the musculoskeletal and hepatic system. Importantly, we showed that the association between physical health and depression and anxiety is partially mediated by individual differences in brain structure. Given physical health assessments preceded brain imaging and mental health questionaries, our results suggest that poor physical health across multiple organ systems, particularly of musculoskeletal, immune, metabolic and hepatic system may lead to subsequent alterations in brain structure, including reductions in gray matter volume and changes in the microstructure of white matter tracts. These structural changes of the brain may lead to or exacerbate symptoms of depression and anxiety as well as neuroticism.

Despite the consistent involvement of global brain structure (i.e., total GM volume, mean FA) in mediating mental health outcomes, the spatial patterns of involved brain regions differ across depression, anxiety and neuroticism. Specifically, brain gray matter volume reductions linking poor physical health and greater depressive symptom severity are widespread. The regions with the strongest mediating effects are located in the orbitofrontal, temporal and insular cortices as well as some parietal regions. These regions are largely consistent with previously reported cortical regions implicated in major depressive disorder ^23,46^. This suggests that regions involved in depression neuropathology may be also susceptible to the negative impact of poor physical health. Whereas the number of regions involved in mediating the relationship between physical health and neuroticism is less than those for depression, most regions were consistently found in orbitofrontal, insula and precentral area. In contrast, brain GM regions linking poor physical health and greater anxiety symptom severity are limited to the middle and superior temporal areas, and to a lesser extent insula, but not in the frontal lobe. This suggests possibly shared neurobiological pathways mediating the physical-mental health relationship between depression, anxiety and neuroticism in temporal and insular cortices, and relatively unique role of orbitofrontal in depression and neuroticism. In terms of the white matter, we found largely shared sets of tracts involved in the pathways leading to depression, anxiety and neuroticism. However, the superior fronto-occipital fasciculus had the strongest mediating effect for depression, whereas the anterior corona radiata and the posterior thalamic radiation had the largest effect on anxiety and neuroticism, respectively. Of note, traits of depression, anxiety and neuroticism are inherently correlated and fluctuate overtime. The relatively distinct findings relating to anxiety may therefore be biased by the discrepancy in the assessment time between anxiety symptoms and the brain as well as the other two mental health measures. Further research is needed to validate the mediating effect of the brain in linking physical health and anxiety.

Our investigation into lifestyle and environmental factors provides a biological understanding of pathways explaining epidemiological observations linking lifestyle with mental health risk and treatment outcomes ^44^. We showed that the impact of lifestyle on mental health, particularly of depression and neuroticism, is partially mediated by the physical health of multiple organs and brain structure. This indicates that although there is a direct effect from lifestyle to mental health, lifestyle factors can influence mental health via influencing the underlying physiology and the brain. This suggests the benefit of personalizing lifestyle-based intervention strategies to suit a person’s physical health and neurobiology. More importantly, we showed that some lifestyle factors, such as physical activity, sleep and diet influence mental health via influencing the physiological function of multiple organ and brain systems. Behavioral interventions focusing on these factors are thus likely to yield improved mental health outcome for most individuals. In contrast, limiting tobacco smoking and alcohol intake are likely beneficial for individuals who have compromised metabolic and pulmonary function, and musculoskeletal and hepatic function, respectively. Indeed, most lifestyle-changing programs in psychiatry primarily focus on physical activity; however, with limited success, ^47^ while the efficacy data available for other lifestyle factors are scarce ^44^. Integrated interventions designed to improve both physical and mental health may yield added health benefits. Further work is needed to determine whether interventions informed by the pathways revealed here would lead to improved physical and mental health and reduce the risk and adverse impact of physical-mental comorbidity.

This study has several limitations. First, brain imaging and mental health assessments were not available at the first study wave of the UK Biobank, when physical health was assessed. We were thus unable to assess whether individual variation in brain structure and mental health symptom severity observed at follow-up is consistent with alterations in brain structure and mental health status assessed longitudinally, consequent to physical health changes. Second, because of the sequential and non-randomized participant assessment, we were unable to assess pathways, whereby poor mental health may lead to poor physical health via influencing brain structure, or alterations in brain structure may lead to poor mental health via influencing physical health. Third, the extent to which our findings are specific to the mental disorders considered requires further investigation, given the high co-occurrence of depression and anxiety. Supplementary analyses were undertaken using a general summary measure of mental health to address this limitation. Fourth, the UK Biobank cohort comprises predominantly white British participants. Further work is needed to assess the generalizability of our findings in a diversity of ethnicities and socioeconomic backgrounds. This is important because the greatest physical-mental health inequalities often persist in low-income countries where evidence-based interventions are most lacking^48^. Finally, this study focuses on brain structure, future work using other neuroimaging modalities such as functional MRI and MR spectroscopy of cerebral metabolites is needed to further characterize the role of the brain in the complex interplay between physical and mental health.

In conclusion, our work provides an integrated model linking physical health, neurobiology and mental health outcomes. Our findings suggest a crucial role of the brain in mediating the relationship between physical and mental health, which is an important step toward bridging the mind-body dualism. The modifiable lifestyle factors identified here can potentially inform the development of targeted interventions to improve both physical and mental health synergistically.

## Methods

### Participants

Physical, physiological, blood- and urine-derived phenotypes and neuroimaging data were sourced from the UK Biobank. The UK Biobank is a large-scale biomedical database and research resource containing genetic, lifestyle and health information from approximately 500,000 participants^35,36^. The UK Biobank has approval from the North West Multi-centre Research Ethics Committee (MREC) to obtain and disseminate data and samples from the participants (http://www.ukbiobank.ac.uk/ethics/). Written informed consent was obtained from all participants.

A subset of the UK Biobank cohort (n=18,083, 7,455 males) were analyzed in the present study (project ID 60698). They were aged 40-70 years (mean 53.7 ± 7.3) at the time of recruitment (2006-2010) and underwent extensive physical and physiological assessments, blood and urine sample assays and questionnaires for environmental and lifestyle factors. Multimodal brain imaging was acquired 4-14 years later (2014-2020) at the age of 45-83 years (mean 62.7 ± 7.5). Among these individuals, 7,749 individuals (3,774 males) had no clinically diagnosed major medical and mental conditions; 10,334 (3,659 males) individuals had a lifetime diagnosis of one of the 4 following common mental disorders including schizophrenia (n=67), bipolar disorder (n=592), depression (n=9,817) and generalized anxiety disorder (n=2,041). The diagnostic and medical information were obtained through self-report (UK Biobank Field ID: 20002) and data linkage to health care records from the UK National Health Services (Field IDs: 41270; 41271; 42040). Details of data curation and diagnostic codes for the four diagnostic groups were described elsewhere ^32^.

### Organ health score

An organ health score was estimated for each organ system and for each individual. The methodology for computing an organ health score was described previously ^32^. In brief, the approach involves two main steps. The first step is to establish an age- and sex-specific normative reference ranges (median and centiles) for each body phenotype derived from physical and physiological assessments and blood and urine assays, using generalized additive models for location, scale and shape (GAMLSS)^49^. A total of 73 body phenotypes were included in this analysis and were grouped into 7 organ systems (cardiovascular, pulmonary, musculoskeletal, immune, renal, hepatic and metabolic) based on its known functional relevance (Supplementary Table 2). A standardized deviation score was computed for each phenotype and for each individual, which quantifies the number of standard deviation an individual deviated from the normative reference median ^37^. The second step is to summarize phenotypic deviation scores at an organ system level, to yield a single health score for each organ and for each individual. The organ health score is a weighted sum of deviation scores (z-scores) across all body phenotypes pertaining to a specific organ system. The weights were estimated according to the importance of each phenotype in differentiating healthy individuals from a patient group that i) was diagnosed with one or more chronic illnesses that primarily affects the organ under consideration; and ii) had no comorbid mental illness. An organ health score quantifies the extent to which an individual’s organ health and function deviates from an age- and sex-specific population median. Details are provided in Tian and colleagues ^32^.

### MRI-derived brain phenotypes

Regional cortical (Desikan-Killiany atlas ^50^) and subcortical GM volume and the mean FA of skeletonized WM tracts (JHU ICBM-DTI-81 atlas ^51^) derived from T1-weighted and diffusion MRI were sourced from the UK Biobank ^36^. The image processing pipeline, artefact removal, cross-modality and cross-individual image alignment, quality control and phenotype estimation are described in detail elsewhere ^52^ and are available in the UK Biobank brain imaging documentation (https://biobank.ctsu.ox.ac.uk/showcase/showcase/docs/brain_mri.pdf). Regional measures were averaged across the left and right hemispheres, resulting in 33 gray and 27 white matter regional metrics (Supplementary Table 3). A standardized deviation score was calculated for each of the regional metrics and two global measures (i.e., total GM volume, mean FA across all WM tracts) for each individual based on established normative reference ranges described previously ^32^.

### Structural equation modeling (path analysis)

SEM was first used to test the significance of pathways (*a*×*b*, Figure 1b) from physical health of organ systems (exogenous variable) to mental health (outcome) via the brain (mediator). A SEM was separately formulated for each organ system and each brain tissue type (GM & WM) and for depression, anxiety and neuroticism, respectively. For paths deemed statistically significant, we next tested the significance of pathways (*a*×*b*×*d*, Figure 1c) from lifestyle factors (exogenous) to mental health (outcome) via physical health (mediator) and the brain (mediator). Similarly, a SEM was separately formulated for each lifestyle factor. The *lavaan* package (version 0.6-16) ^53^ in R for SEM was used to estimate the regression coefficient of each path. All variables were normalized by mean and standard deviation across all individuals before model fitting to ensure that the magnitude of regression coefficient is comparable across models. The model was repeatedly fitted for 5,000 bootstrapped samples to estimate the confidence intervals. The p-value of each path was given by the proportion of bootstrapped samples where the estimated coefficient is larger (positive effect) or smaller (negative effect) than zero. The false discovery rate was further controlled at 0.05 across the set of mediation effects tested. The goodness of model fit of each SEM model was evaluated using the following three indices: the comparative fit index (CTI), the root mean square error of approximation (RMSEA) and the standardized root mean square residual (SRMR), as recommended by Kline ^54^. The chi-square statistic was not considered due to its sensitivity to sample size. A model was deemed satisfactory if CFI>0.90 or RMSEA<0.05 or SRMR<0.08 ^55^.

### Summary measure of mental health outcomes

Principal component analysis was used to derive a general summary measure of mental health status across depression, anxiety and neuroticism. The first principal component explained 68.8% variance and was used as the summary measure in supplementary analyses.

## Acknowledgments

This research has been conducted using data from UK Biobank (https://www.ukbiobank.ac.uk/), a major biomedical database. We that the UK Biobank for making the data available, and to all study participants, who generously donated their time to make this resource possible. YET was supported by a Mary Lugton Postdoctoral Fellowship. AZ was supported by a Senior Rebecca L. Cooper Fellowship. ETB was supported by a Senior Investigator award from the National Institute of Health Research (UK).

## Author Contributions

YET and AZ conceived the idea and designed the study. YET compiled the data, performed the analyses, prepared the visualizations and drafted the manuscript. JHC and ETB provided critical conceptual input. AZ supervised the research. All authors provided critical feedback and editing of the final manuscript.

## Data Availability

Data were obtained from the UK Biobank. Researchers can register to access all data used in this study via the UK Biobank Access Management System (https://bbams.ndph.ox.ac.uk/ams/).

## Code availability

Matlab code (R2022b, Natick, Massachusetts: The MathWorks Inc.) for computing organ health scores is available on GitHub (https://github.com/yetianmed/OrganHealthScore). Structural equation modeling was performed using the *lavaan* package (version 0.6-16) in R.

---

**Figure 1.**
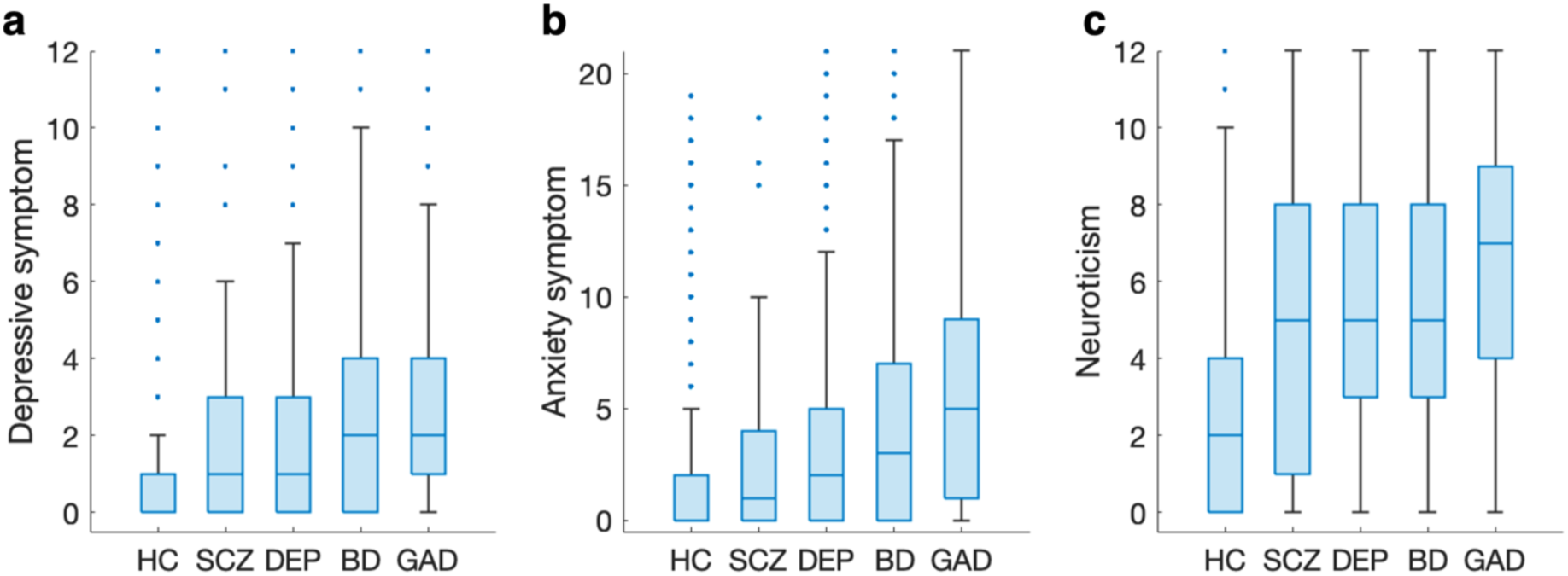
Mental health assessment. The severity of depressive (**a**), anxiety (**b**) and neuroticism (**c**) symptoms in healthy comparison (HC) individuals and individuals who had a lifetime diagnosis of one of the following 4 mental disorders: schizophrenia (SCZ), depression (DEP), bipolar disorder (BD) and generalized anxiety disorder (GAD). Depressive symptom severity was represented by the total score of the Recent Depressive Symptom (RDS-4) ^1^. The score was re-scaled so that a score of zero indicates no recent depressive symptoms. Anxiety symptom severity was represented by the total score of the Generalized Anxiety Disorder (GAD-7) scale ^2^. Neuroticism was assessed by the total score of the Eysenck Neuroticism (N-12) questionnaire ^3^.

**Figure 2.**
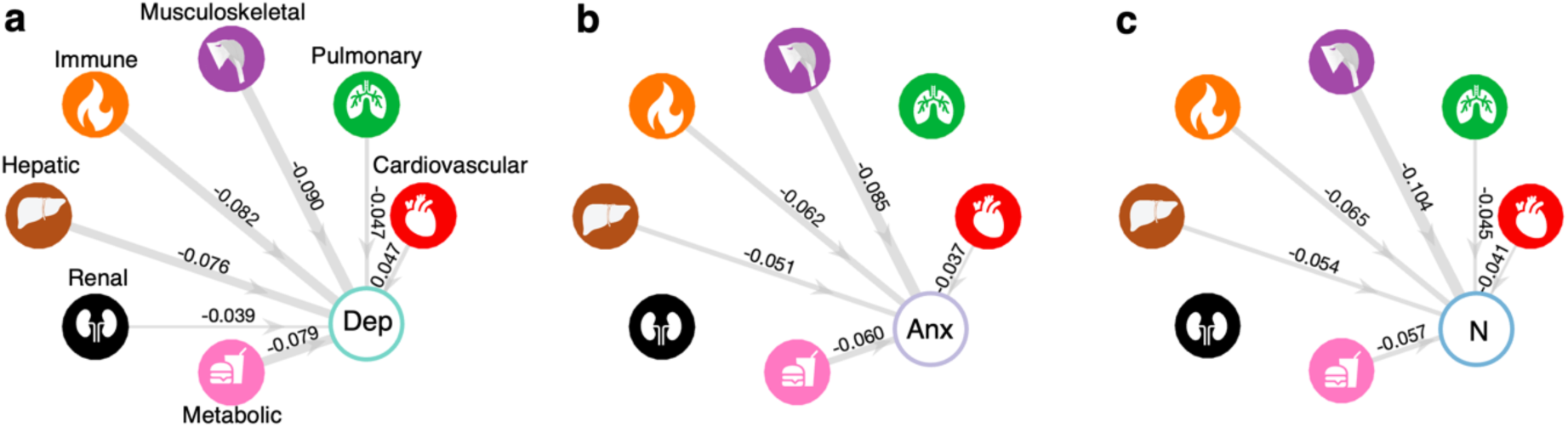
Associations between organ health and mental health. Prospective associations between baseline organ health of 7 body systems and the severity of depression (**a**), anxiety (**b**) and neuroticism (**c**) symptoms at follow-up. Links are shown for significant associations between the health score of organ systems and mental health measures, adjusting for sex and age at baseline (p<0.05, two-tailed, false discovery rate (FDR) corrected across 7 organs). Edge thickness reflects standardized regression coefficients (*β*). Dep, depression; Anx, anxiety; N, neuroticism.

**Figure 3.**
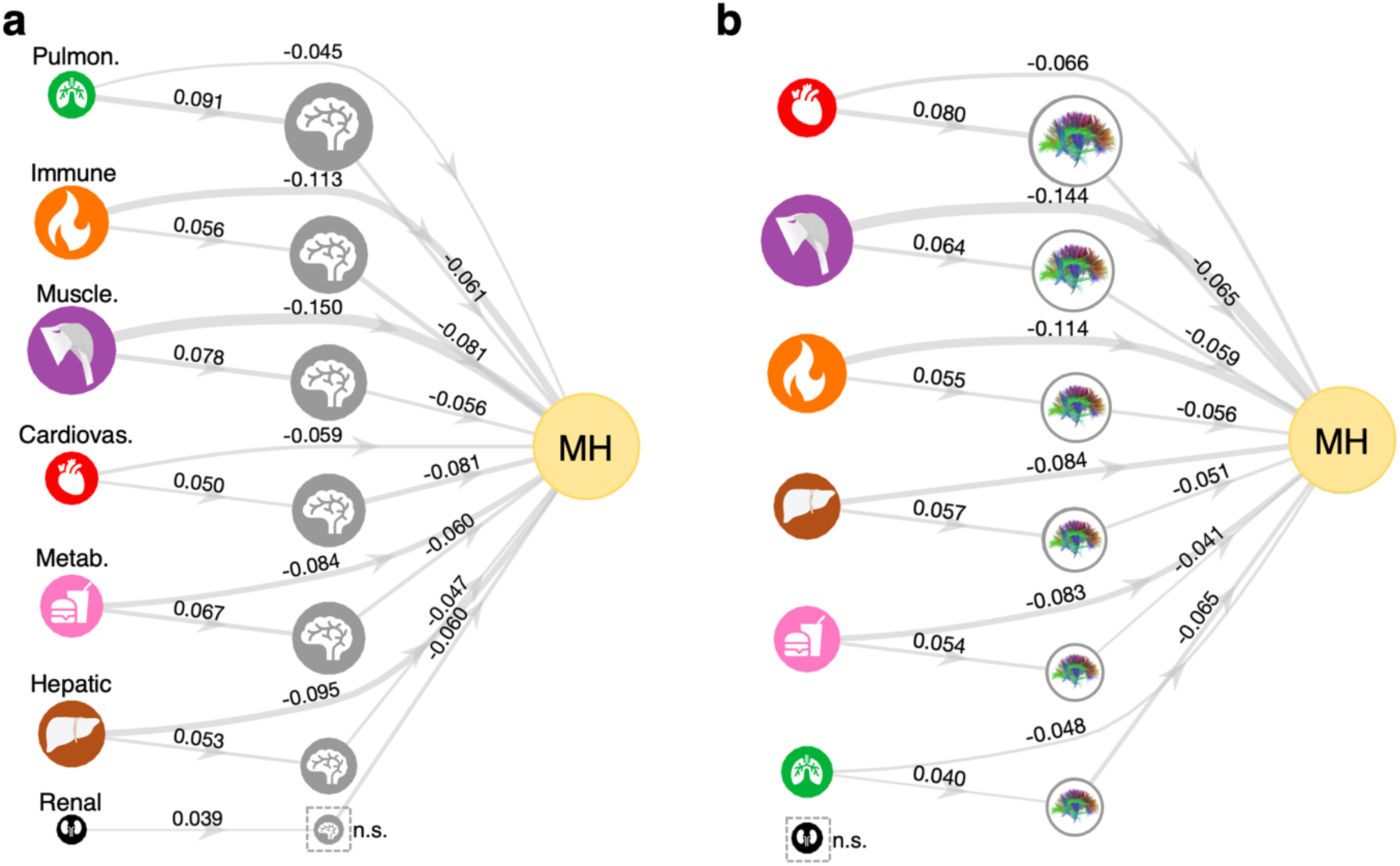
Mediating effects of gray and white matter on physical-mental health associations. Pathways linking organ health, brain gray matter volume (**a**)/ fractional anisotropy (FA) of white matter (**b**), and mental health. A structural equation model (SEM) linking the health of each organ system to a summary measure of mental health (MH) across depression, anxiety and neuroticism. The summary measure was indicated by the first principal component of the three measures. Links are shown for significant paths linking organ health (exogeneous) and mental health (outcome) via brain gray matter volume (**a**) or white matter tracts (**b**, mediator) inferred from SEM (false discovery rate corrected for 7 organs). Node size of organs is modulated by the direct effect from organs to mental health outcome. Node size of the brain is modulated by its mediating effect and ranked in decreasing order. Edge thickness reflects regression coefficients estimated for edges comprising the SEM. Non-significant mediating effect of the brain is indicated by a dashed square. Pulmon., pulmonary; Muscle., musculoskeletal; Cardiovas., cardiovascular; Dep, depression; Anx, anxiety; N, neuroticism; n.s., non-significant.

**Figure 4.**
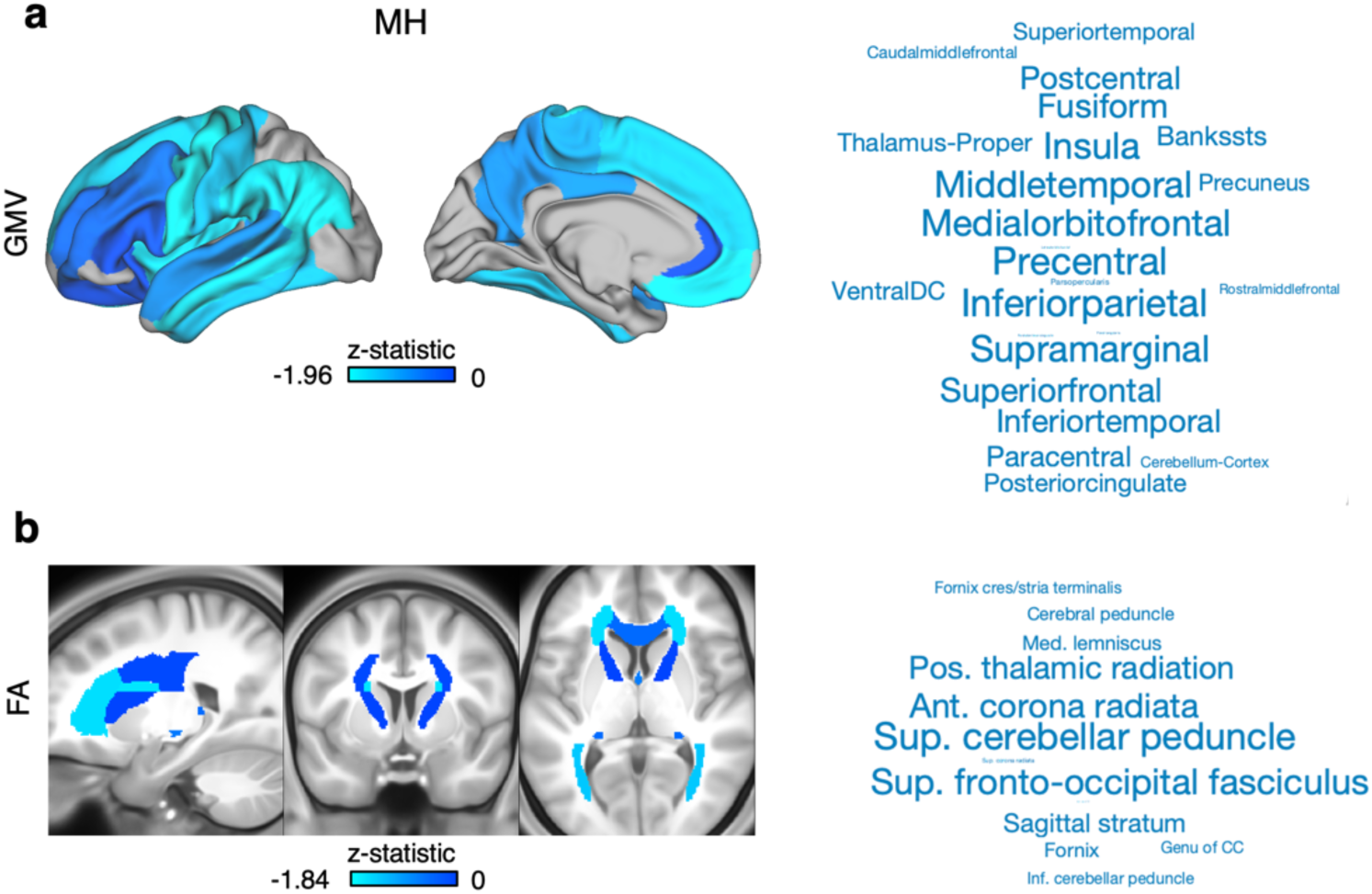
Mediating effects of gray and white matter regions on physical-mental health associations. A structural equation model (SEM) was fitted for each gray matter region, white matter tract and for each organ system. Significant mediating effects (z-statistic) of brain regions (p<0.05, FDR-corrected for 33 gray and 27 white matter regions) were averaged across 7 organ systems to provide a consensus mediating effect map for overall mental health. The overall mental health (MH) score was indicated by the first principal component across three measures, i.e., depression, anxiety and neuroticism. The average z-statistic for cortical gray matter volume (Desikan-Killianny atlas) are rendered on cortical surface for visualization. Word clouds show top-ranked cortical and subcortical regions. The font size is scaled according to the absolute value of the average z-statistic. Similarly, the average z-statistic for regional fractional anisotropy (JHU ICBM-DTI-81 atlas) are rendered in standard Montreal Neurological Institute (MNI)-152 anatomical space. Word clouds show top-ranked white matter tracts. The font size is scaled according to the absolute value of the average z-statistic.

**Figure 5.**
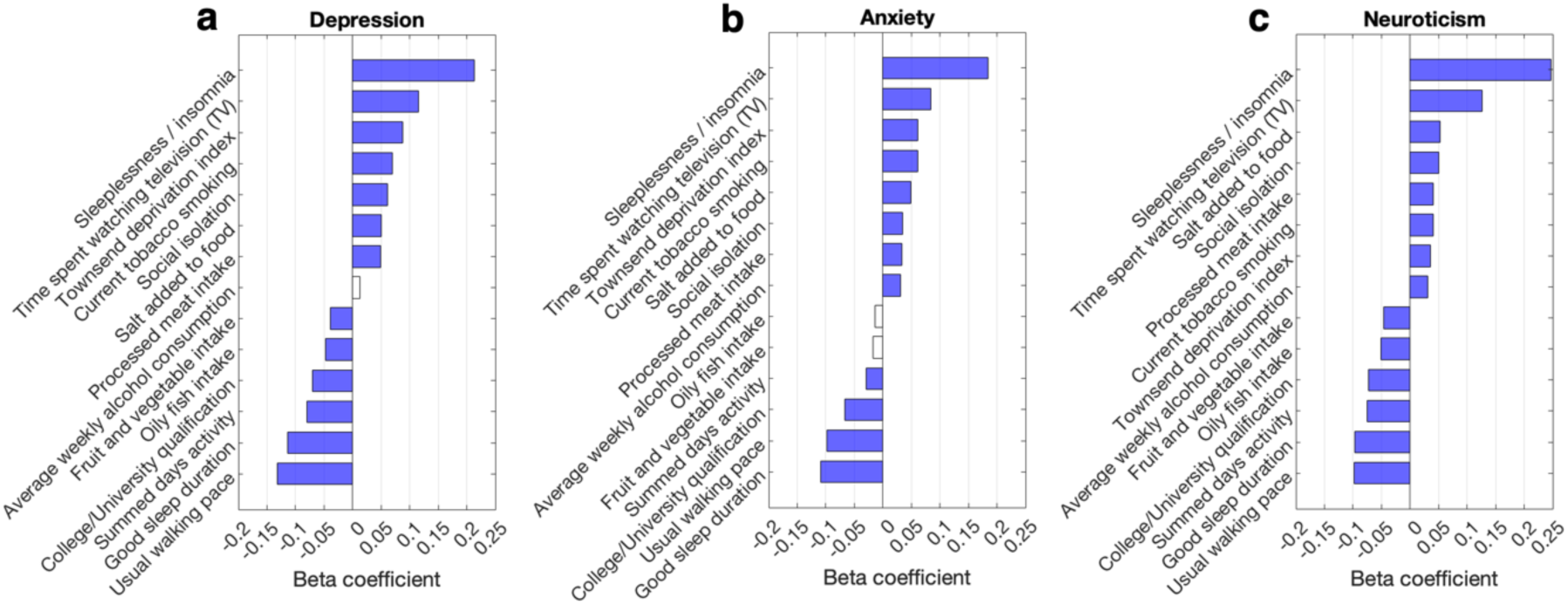
Associations between lifestyle factors and mental health. Bar plots show the prospective associations between baseline exposure to lifestyle factors and the severity of depression (**a**), anxiety (**b**) and neuroticism (**c**) symptoms at follow-up. Colored bars indicate lifestyle factors with significant associations, adjusting for sex and age at baseline (p<0.05, FDR corrected across 14 lifestyle factors). Lifestyle factors are ordered from top to bottom according to decreasing regression coefficient.

**Table 1.** Baseline demographic characteristics.

| **Group** | **No. of individuals** | **Age, mean (SD), year** | **Male, No. (%)** |
| --- | --- | --- | --- |
| Healthy comparison | 7,749 | 53.1 (7.3) | 3,774 (48.7) |
| Schizophrenia | 67 | 54.8 (7.4) | 41 (61.2) |
| Bipolar disorder | 592 | 53.8 (7.4) | 287 (48.5) |
| Depression | 9,817 | 54.2 (7.3) | 3,397 (34.7) |
| Generalized anxiety disorder | 2,041 | 53.4 (7.2) | 751 (35.4) |

**Table 2.**
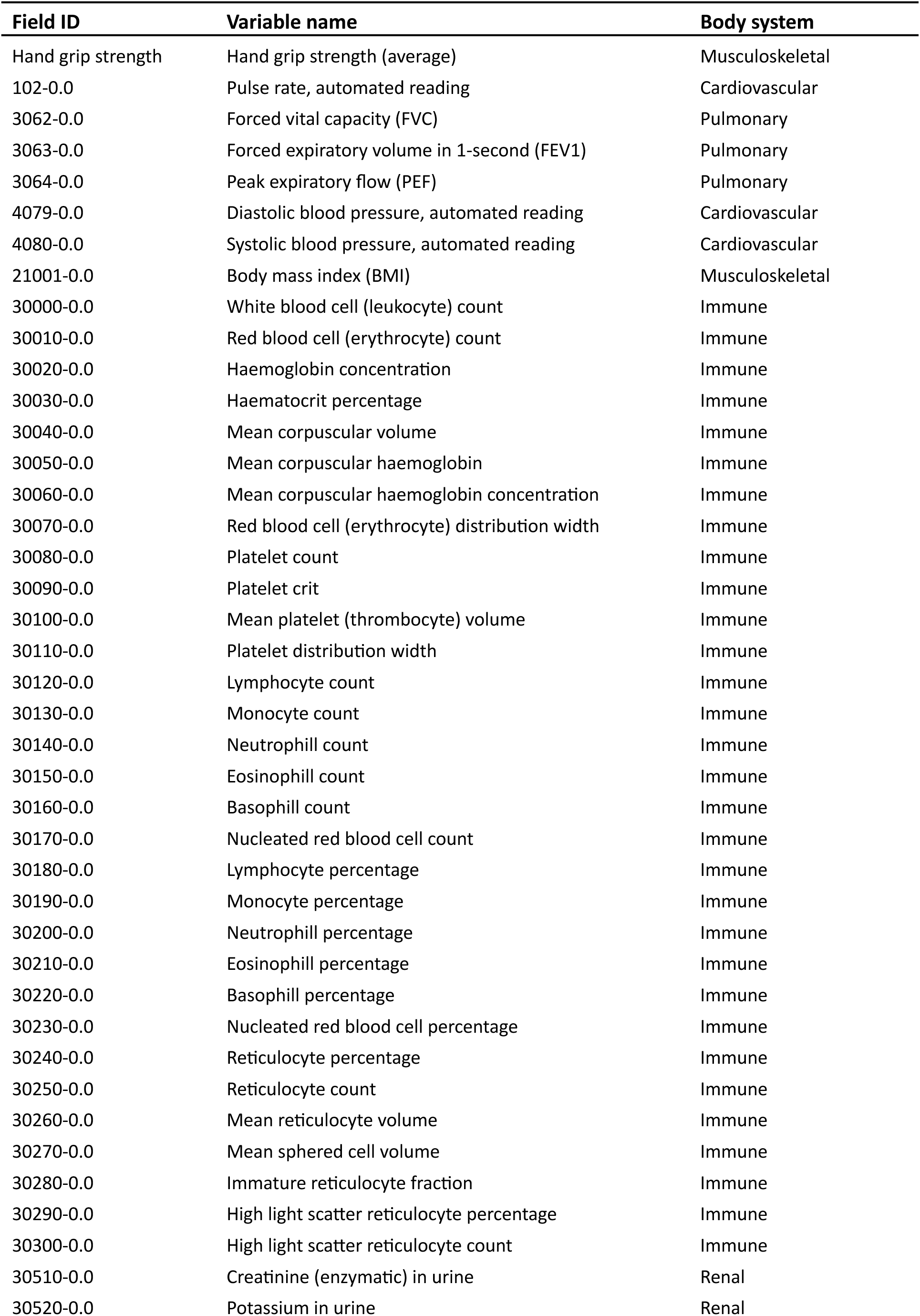
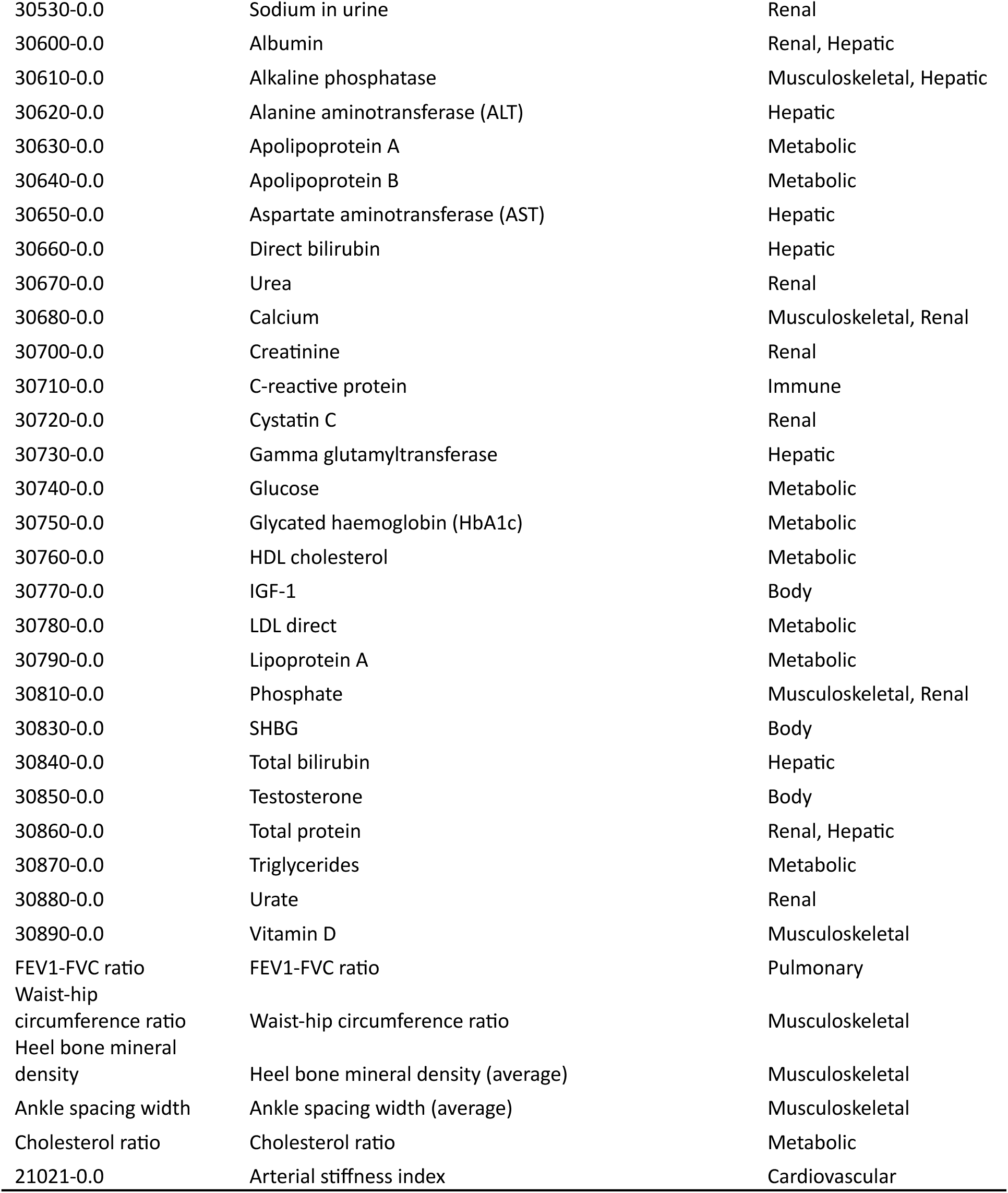
Body phenotypes.

| Field ID | Variable name | Body system |
| --- | --- | --- |
| Hand grip strength | Hand grip strength (average) | Musculoskeletal |
| 102-0.0 | Pulse rate, automated reading | Cardiovascular |
| 3062-0.0 | Forced vital capacity (FVC) | Pulmonary |
| 3063-0.0 | Forced expiratory volume in 1-second (FEV1) | Pulmonary |
| 3064-0.0 | Peak expiratory flow (PEF) | Pulmonary |
| 4079-0.0 | Diastolic blood pressure, automated reading | Cardiovascular |
| 4080-0.0 | Systolic blood pressure, automated reading | Cardiovascular |
| 21001-0.0 | Body mass index (BMI) | Musculoskeletal |
| 30000-0.0 | White blood cell (leukocyte) count | Immune |
| 30010-0.0 | Red blood cell (erythrocyte) count | Immune |
| 30020-0.0 | Haemoglobin concentration | Immune |
| 30030-0.0 | Haematocrit percentage | Immune |
| 30040-0.0 | Mean corpuscular volume | Immune |
| 30050-0.0 | Mean corpuscular haemoglobin | Immune |
| 30060-0.0 | Mean corpuscular haemoglobin concentration | Immune |
| 30070-0.0 | Red blood cell (erythrocyte) distribution width | Immune |
| 30080-0.0 | Platelet count | Immune |
| 30090-0.0 | Platelet crit | Immune |
| 30100-0.0 | Mean platelet (thrombocyte) volume | Immune |
| 30110-0.0 | Platelet distribution width | Immune |
| 30120-0.0 | Lymphocyte count | Immune |
| 30130-0.0 | Monocyte count | Immune |
| 30140-0.0 | Neutrophill count | Immune |
| 30150-0.0 | Eosinophill count | Immune |
| 30160-0.0 | Basophill count | Immune |
| 30170-0.0 | Nucleated red blood cell count | Immune |
| 30180-0.0 | Lymphocyte percentage | Immune |
| 30190-0.0 | Monocyte percentage | Immune |
| 30200-0.0 | Neutrophill percentage | Immune |
| 30210-0.0 | Eosinophill percentage | Immune |
| 30220-0.0 | Basophill percentage | Immune |
| 30230-0.0 | Nucleated red blood cell percentage | Immune |
| 30240-0.0 | Reticulocyte percentage | Immune |
| 30250-0.0 | Reticulocyte count | Immune |
| 30260-0.0 | Mean reticulocyte volume | Immune |
| 30270-0.0 | Mean sphered cell volume | Immune |
| 30280-0.0 | Immature reticulocyte fraction | Immune |
| 30290-0.0 | High light scatter reticulocyte percentage | Immune |
| 30300-0.0 | High light scatter reticulocyte count | Immune |
| 30510-0.0 | Creatinine (enzymatic) in urine | Renal |
| 30520-0.0 | Potassium in urine | Renal |
| 30530-0.0 | Sodium in urine | Renal |
| 30600-0.0 | Albumin | Renal, Hepatic |
| 30610-0.0 | Alkaline phosphatase | Musculoskeletal, Hepatic |
| 30620-0.0 | Alanine aminotransferase (ALT) | Hepatic |
| 30630-0.0 | Apolipoprotein A | Metabolic |
| 30640-0.0 | Apolipoprotein B | Metabolic |
| 30650-0.0 | Aspartate aminotransferase (AST) | Hepatic |
| 30660-0.0 | Direct bilirubin | Hepatic |
| 30670-0.0 | Urea | Renal |
| 30680-0.0 | Calcium | Musculoskeletal, Renal |
| 30700-0.0 | Creatinine | Renal |
| 30710-0.0 | C-reactive protein | Immune |
| 30720-0.0 | Cystatin C | Renal |
| 30730-0.0 | Gamma glutamyltransferase | Hepatic |
| 30740-0.0 | Glucose | Metabolic |
| 30750-0.0 | Glycated haemoglobin (HbA1c) | Metabolic |
| 30760-0.0 | HDL cholesterol | Metabolic |
| 30770-0.0 | IGF-1 | Body |
| 30780-0.0 | LDL direct | Metabolic |
| 30790-0.0 | Lipoprotein A | Metabolic |
| 30810-0.0 | Phosphate | Musculoskeletal, Renal |
| 30830-0.0 | SHBG | Body |
| 30840-0.0 | Total bilirubin | Hepatic |
| 30850-0.0 | Testosterone | Body |
| 30860-0.0 | Total protein | Renal, Hepatic |
| 30870-0.0 | Triglycerides | Metabolic |
| 30880-0.0 | Urate | Renal |
| 30890-0.0 | Vitamin D | Musculoskeletal |
| FEV1-FVC ratio | FEV1-FVC ratio | Pulmonary |
| Waist-hip circumference ratio | Waist-hip circumference ratio | Musculoskeletal |
| Heel bone mineral density | Heel bone mineral density (average) | Musculoskeletal |
| Ankle spacing width | Ankle spacing width (average) | Musculoskeletal |
| Cholesterol ratio | Cholesterol ratio | Metabolic |
| 21021-0.0 | Arterial stiffness index | Cardiovascular |

**Table 3.**
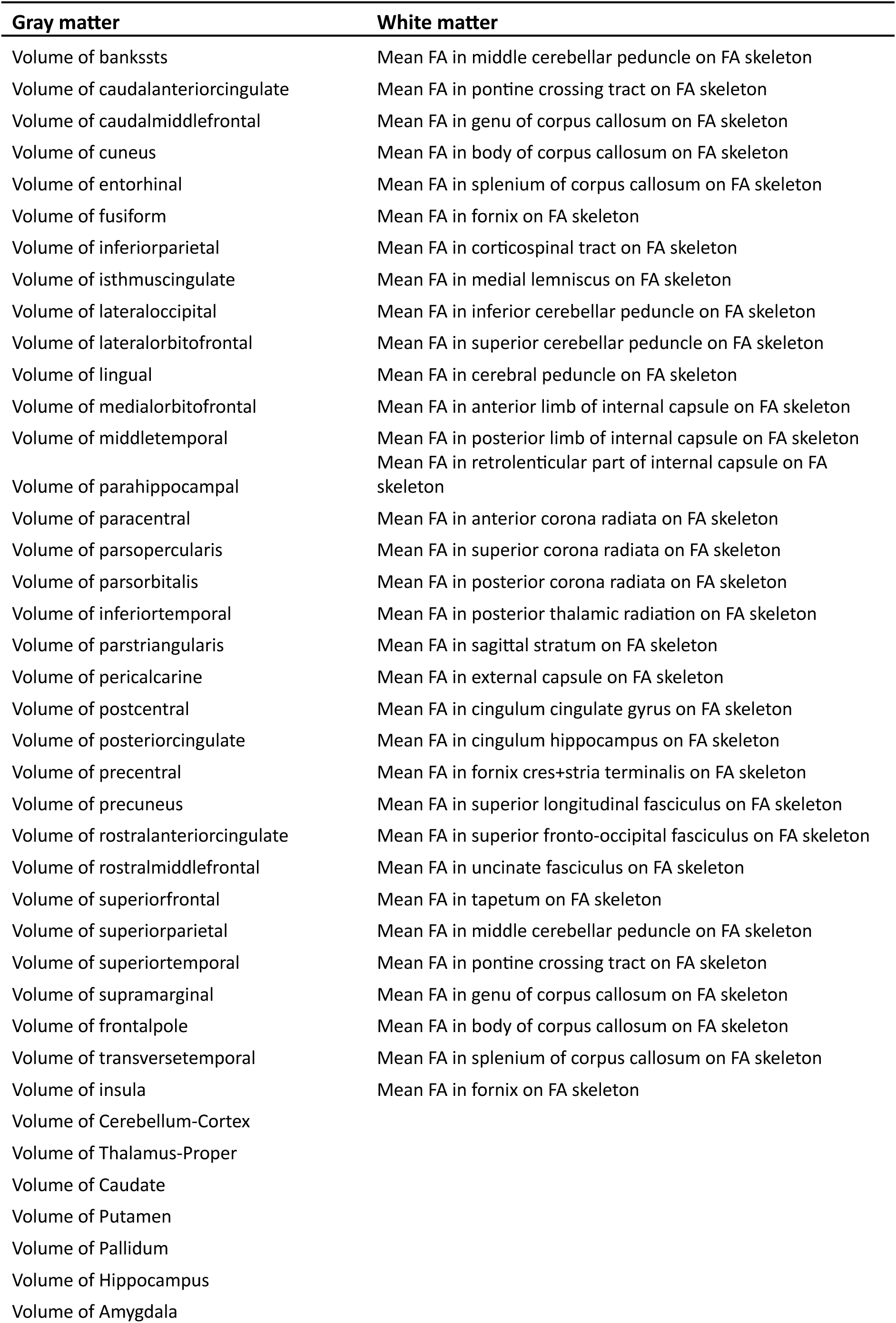
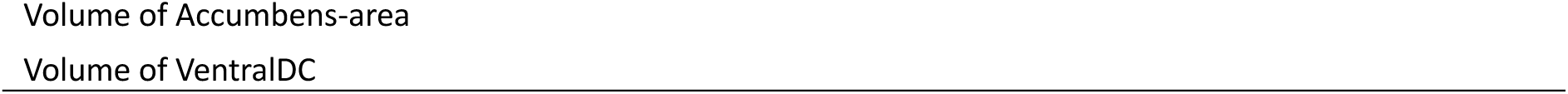
Brain phenotypes.

